# Systematic evaluation of transcriptomic disease risk and diagnostic biomarker overlap between COVID-19 and tuberculosis: a patient-level meta-analysis

**DOI:** 10.1101/2020.11.25.20236646

**Authors:** Dylan Sheerin, Abhimanyu, Xutao Wang, W Evan Johnson, Anna Coussens

**Affiliations:** Infectious Diseases and Immune Defence Division, The Walter & Eliza Hall Institute of Medical Research, Parkville 3279, VIC, Australia; Wellcome Centre for Infectious Diseases in Africa, Institute of Infectious Disease and Molecular Medicine, University of Cape Town, Anzio Rd, Observatory, 7925, Western Cape, South Africa; Division of Computational Biomedicine and Bioinformatics Program, Boston University, Boston, MA, USA; Department of Biostatistics, Boston University, Boston, MA, USA; Department of Medical Biology, University of Melbourne, Parkville 3010, VIC, Australia

## Abstract

**Background:** The novel coronavirus, SARS-CoV-2, has increased the burden on healthcare systems already strained by a high incidence of tuberculosis (TB) as co-infection and dual presentation are occurring in syndemic settings. We aimed to understand the interaction between these diseases by profiling COVID-19 gene expression signatures on RNA-sequencing data from TB-infected individuals.

**Methods:** We performed a systematic review and patient-level meta-analysis by querying PubMed and pre-print servers to derive eligible COVID-19 gene expression signatures from human whole blood (WB), PBMCs or BALF studies. A WB influenza dataset served as a control respiratory disease signature. Three large TB RNA-seq datasets, comprising multiple cohorts from the UK and Africa and consisting of TB patients across the disease spectrum, were chosen to profile these signatures. Putative “COVID-19 risk scores” were generated for each sample in the TB datasets using the TBSignatureProfiler package. Risk was stratified by time to TB diagnosis in progressors and contacts of pulmonary and extra-pulmonary TB. An integrative analysis between TB and COVID-19 single-cell RNA-seq data was performed and a population-level meta-analysis was conducted to identify shared gene ontologies between the diseases and their relative enrichment in COVID-19 disease severity states.

**Results:** 35 COVID-19 gene signatures from nine eligible studies comprising 98 samples were profiled on TB RNA-seq data from 1181 samples from 853 individuals. 25 signatures had significantly higher COVID-19 risk in active TB (ATB) compared with latent TB infection (p <0·005), 13 of which were validated in two independent datasets. *FCN1*- and *SPP1*-expressing macrophages enriched in BALF during severe COVID-19 were identified in circulation during ATB. Shared perturbed ontologies included antigen presentation, epigenetic regulation, platelet activation, and ROS/RNS production were enriched with increasing COVID-19 severity. Finally, we demonstrate that the overlapping transcriptional responses may complicate development of blood-based diagnostic signatures of co-infection.

**Interpretation:** Our results identify shared dysregulation of immune responses in COVID-19 and TB as a dual risk posed by co-infection to COVID-19 severity and TB disease progression. These individuals should be followed up for TB in the months subsequent to SARS-CoV-2 diagnosis.

## Background

Three months after it was first detected, SARS-CoV-2 was declared to have caused the first global pandemic of the 21st Century. In the absence of an effective treatment or vaccine, mortality in the 35 million diagnosed thus far, currently stands at 2.9% (as of October 2020). By comparison tuberculosis (TB), a similar respiratory infection and humanity’s longest continuing pandemic, causes 10 million annual cases, and has a mortality of 12–20%, the upper bound including those HIV-coinfected^1^. This high mortality, of roughly 4000 people a day, exists despite a vaccine that reduces infant mortality and antibiotics which have reduced mortality from the pre-antibiotic era by 50%. With an estimated quarter of the world’s population infected with *Mycobacterium tuberculosis* (*Mtb*), it could be said that TB is a silent killer. Silent in that during the current pandemic, TB has killed roughly the same number of people a day as COVID-19 currently; with the socio-economic and health systems impact of COVD-19 lockdowns estimated to result in an additional 6.8 million TB cases and 1.4 million TB deaths over the next 5 years^2,3^. Countries where TB-HIV already causes high mortality were shielded from previous SARS and MERS outbreaks, and thus the interaction of these coronaviruses with concurrent TB co-infection, has not previously been experienced. COVID-19 and TB share a symptomatic presentation of productive cough, fever and shortness of breath, and clinical parameters of raised C-reactive protein (CRP), erythrocyte sedimentation rate (ESR), D-Dimer, interleukin (IL)-6, leukopenia and neutrophilia. The similarity in clinical parameters and aspects of underlying immunological reactions suggests co-infection will not only complicate diagnostic algorithms, it indicates a potentially fatal convergence in immunopathogenesis. Emerging case studies and population level data indicate TB patients, and those latently infected or with a history of TB are at increased risk of severe COVID-19^4-7^.

In order to develop contextually appropriate treatment and risk mitigation interventions in communities where the potential for *Mtb*-SARS-CoV-2 co-infection is high, we urgently need to understand how the immunopathogenesis of these two respiratory pathogens interact. We therefore conducted a systematic transcriptomic evaluation of whole blood (WB), peripheral blood mononuclear cell (PBMC) and bronchoalveolar lavage fluid (BALF) signatures associated with COVID-19 clinical severity and the spectrum of asymptomatic and symptomatic TB. Our findings suggest that subclinical and active TB (ATB) increase the risk of severe COVID-19 disease, due to increased abundance of circulating myeloid subpopulations also found in the lungs of severe COVID-19 patients. This shared pathway of immunopathogenesis also suggests that SARS-CoV-2 infection will trigger increased progression to TB disease. COVID-19 may therefore pose the biggest threat to ending the TB epidemic since HIV-1 and the modelling increase of 6.8 million extra TB cases in the next 5 years is significantly underestimated^8^.

## Methods

### Search strategy and selection criteria

The hypothesis that WB transcriptomic signatures present in those with existing TB infection will increase risk of severe COVID-19 and interfere in diagnostic biomarker selection was evaluated using a combination of transcriptomic data from COVID-19 patients and WB RNA-seq data from studies of TB disease progression on which the COVID-19 signatures were evaluated. A literature search of published and pre-print manuscripts was conducted on the NIH PubMed and bioRxiv, medRxiv, and SSRN servers uploaded/published between 01/02/20 and 20/09/20 (figure 1). The WB influenza dataset, GSE111368^9^, was used to generate an influenza virus control signature. The curatedTBData package,^10^ which includes 48 publicly available TB RNA-seq datasets, was used to identify eligible TB datasets, that included individuals who progressed to TB during the duration of study follow-up, with RNA-seq data at baseline and time of diagnosis, and patient-level meta-data including time to TB progression. Selection criteria and cohort datasets are described in detail in appendix 1 p2 and p5 (table 1–2).

**Table 1.**
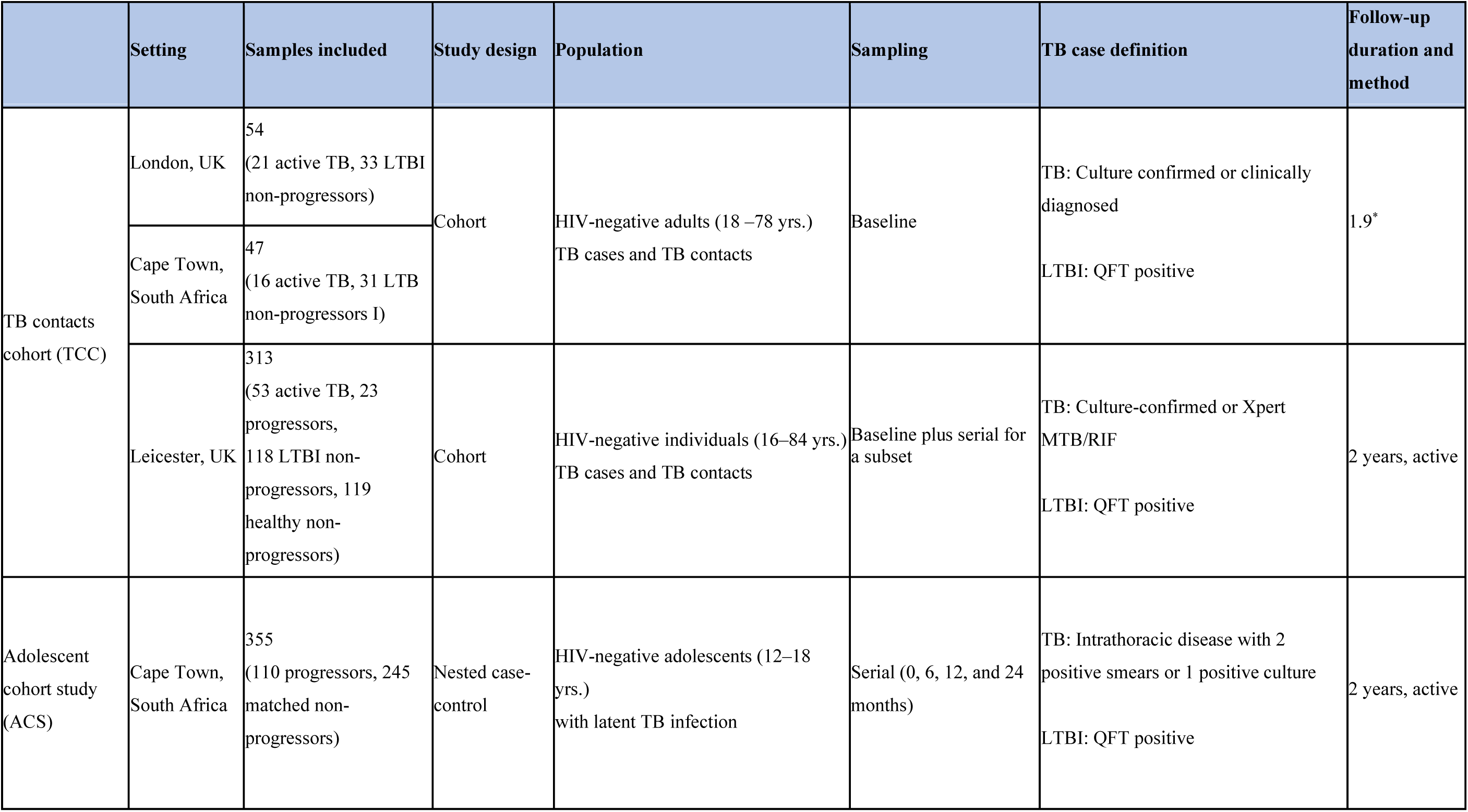
Characteristics of the datasets used to profile COVID-19 risk from COVID-19 immune cell and pathway signatures.

**Table 2.**
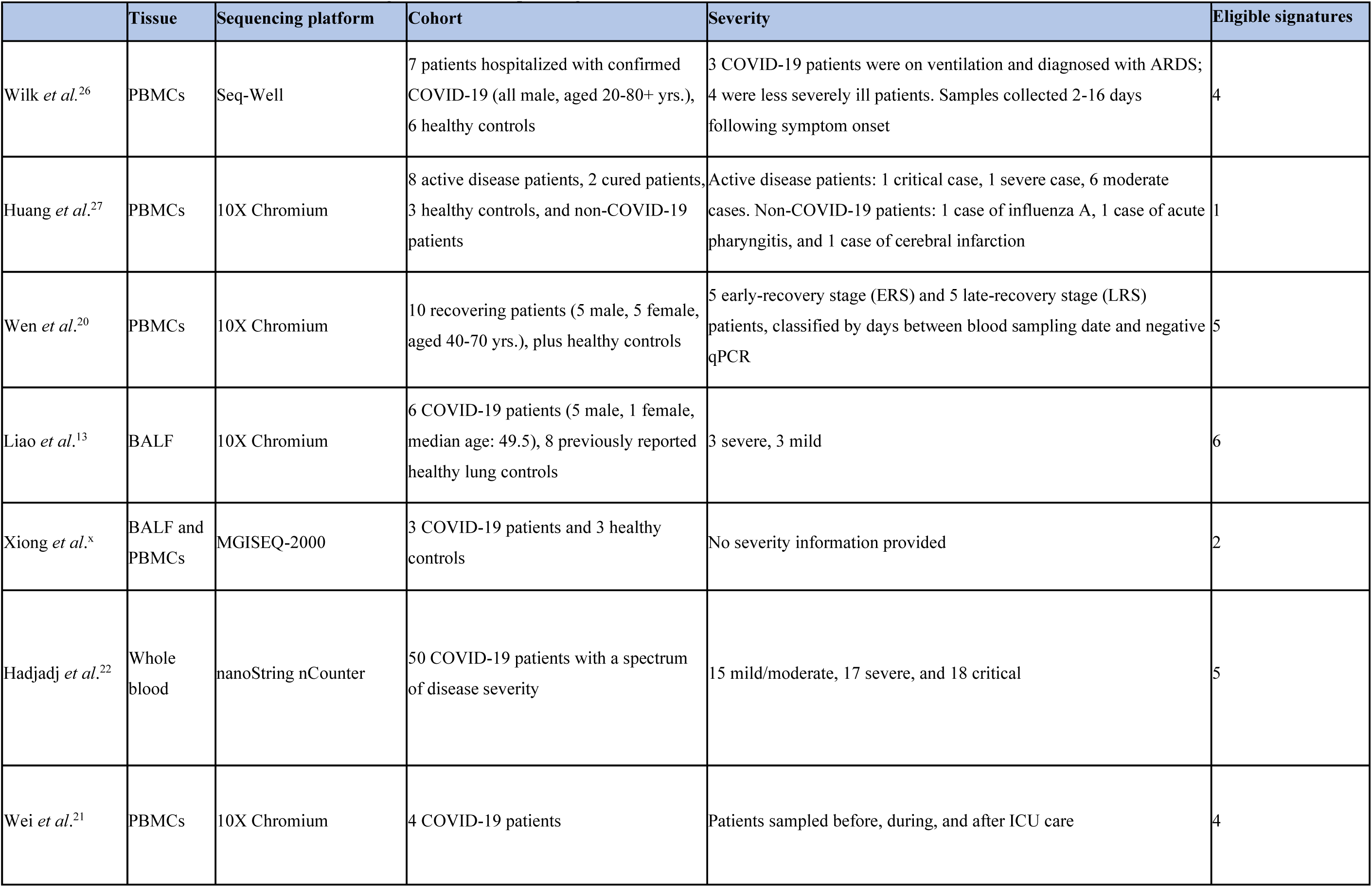
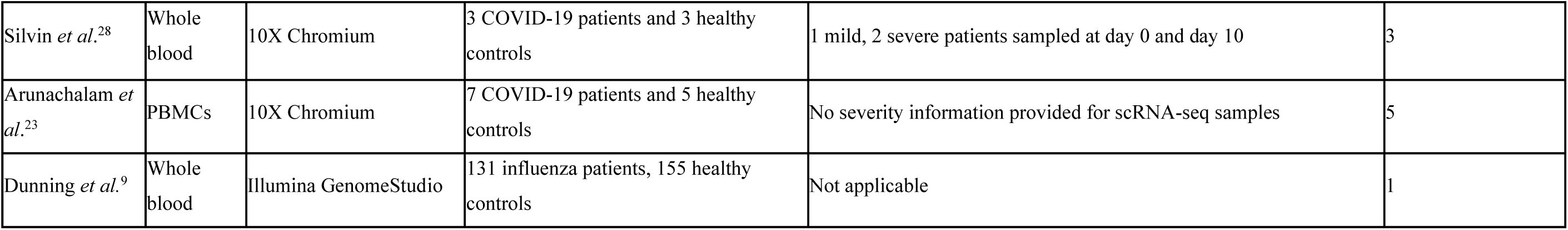
Characteristics of studies used to derive signatures for risk profiling on tuberculosis datasets.

|  | Tissue | Sequencing platform | Cohort | Severity | Eligible signatures |
| --- | --- | --- | --- | --- | --- |
| Wilk *et al.*<sup>26</sup> | PBMCs | Seq-Well | 7 patients hospitalized with confirmed COVID-19 (all male, aged 20-80+ yrs.), 6 healthy controls | 3 COVID-19 patients were on ventilation and diagnosed with ARDS; 4 were less severely ill patients. Samples collected 2-16 days following symptom onset | 4 |
| Huang *et al.*<sup>27</sup> | PBMCs | 10X Chromium | 8 active disease patients, 2 cured patients, 3 healthy controls, and non-COVID-19 patients | Active disease patients: 1 critical case, 1 severe case, 6 moderate cases. Non-COVID-19 patients: 1 case of influenza A, 1 case of acute pharyngitis, and 1 case of cerebral infarction | 1 |
| Wen *et al.*<sup>20</sup> | PBMCs | 10X Chromium | 10 recovering patients (5 male, 5 female, aged 40-70 yrs.), plus healthy controls | 5 early-recovery stage (ERS) and 5 late-recovery stage (LRS) patients, classified by days between blood sampling date and negative qPCR | 5 |
| Liao *et al.*<sup>13</sup> | BALF | 10X Chromium | 6 COVID-19 patients (5 male, 1 female, median age: 49.5), 8 previously reported healthy lung controls | 3 severe, 3 mild | 6 |
| Xiong *et al.*<sup>x</sup> | BALF and PBMCs | MGISEQ-2000 | 3 COVID-19 patients and 3 healthy controls | No severity information provided | 2 |
| Hadjadj *et al.*<sup>22</sup> | Whole blood | nanoString nCounter | 50 COVID-19 patients with a spectrum of disease severity | 15 mild/moderate, 17 severe, and 18 critical | 5 |
| Wei *et al.*<sup>21</sup> | PBMCs | 10X Chromium | 4 COVID-19 patients | Patients sampled before, during, and after ICU care | 4 |
| Silvin *et al.*<sup>28</sup> | Whole blood | 10X Chromium | 3 COVID-19 patients and 3 healthy controls | 1 mild, 2 severe patients sampled at day 0 and day 10 | 3 |
| Arunachalam *et al.*<sup>23</sup> | PBMCs | 10X Chromium | 7 COVID-19 patients and 5 healthy controls | No severity information provided for scRNA-seq samples | 5 |
| Dunning *et al.*<sup>9</sup> | Whole blood | Illumina GenomeStudio | 131 influenza patients, 155 healthy controls | Not applicable | 1 |

**Figure 1.**
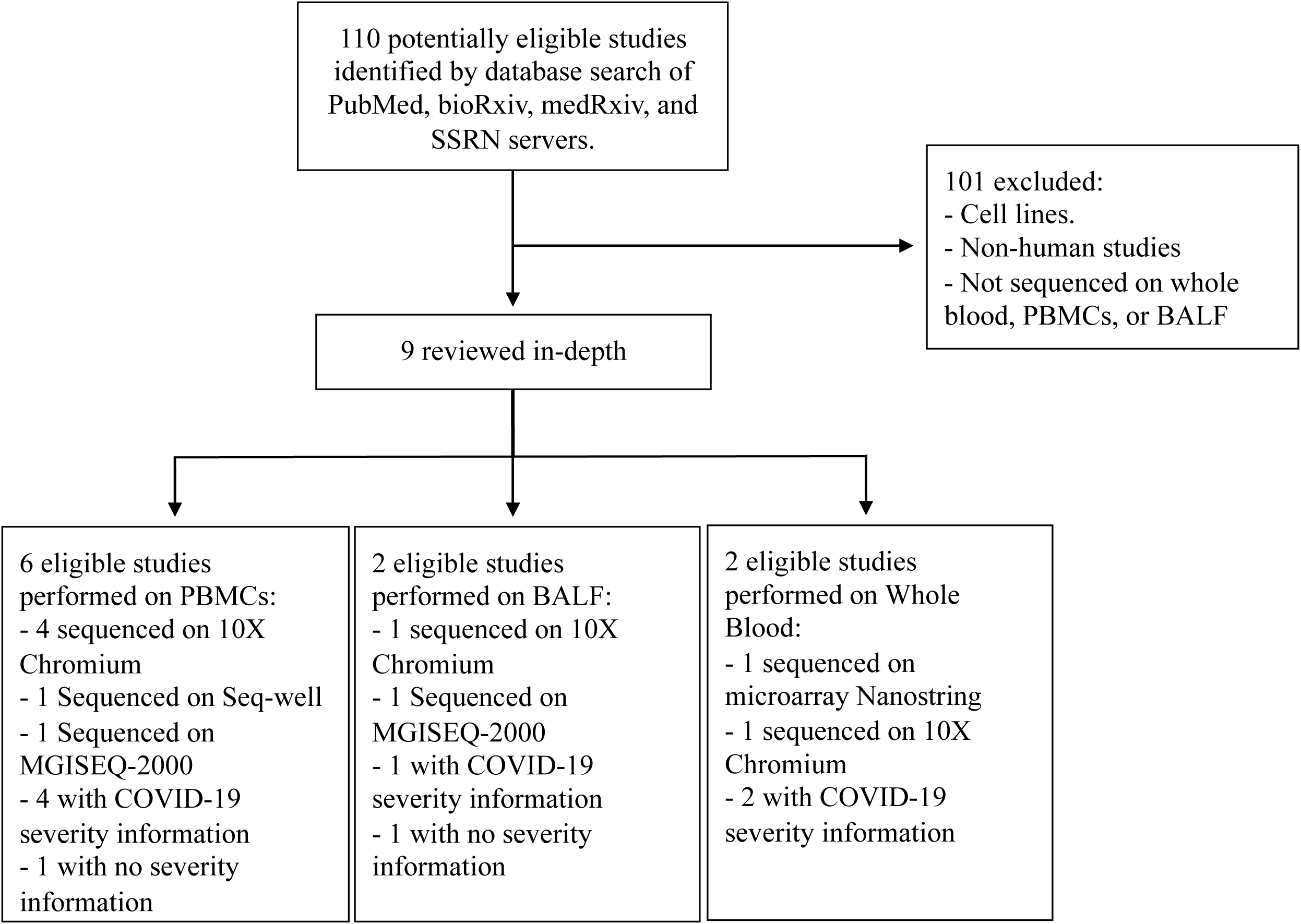
PRISMA flow chart of study selection. PBMCs, peripheral blood mononuclear cells; BALF, bronchoalveolar lavage fluid.

### Data Analysis

TB RNA-seq data were downloaded for eligible studies using the curatedTBData package,^10^ as outlined in appendix 1 p2. Samples collected at all time points were included. The eligible COVID-19 and influenza control signatures (appendix 2 p1) were evaluated independently against the patient-level TB RNA-seq data, generating individual-sample putative “COVID-19 risk score” using gene set variation analysis (GSVA)^11^ with the TBSignatureProfiler package^12^ (appendix 1 p2), and score significance, as compared to latent TB infection (LTBI) controls, calculated by Bonferroni-corrected t-test (appendix 2 p2–4).

Single cell (sc)RNA-seq integrative comparison was conducted using the identified COVID-19 bronchoalveolar lavage fluid (BALF) dataset^13^ downloaded from the NIH GEO database (GSE145926) and a TB PBMC dataset^14^ downloaded from NCBI Short Read Archive (SRA, SRR11038989-SRR11038995). *filtered_feature_bc_matrix*.*h5* files were read into RStudio using the Seurat *Read10X_h5* function (v3.0).^15^ Datasets were independently normalised before the Seurat *FindIntegrationAnchors* function was applied to generate a corrected data matrix for joint analysis. Functional enrichment analysis and cohort selection is described in detail in appendix 1 (p3). Briefly, selected datasets were evaluated using the Metascape^16^ statistically enriched ontology terms, generating enriched pathways (appendix 1 p3–4). Pathway-associated gene clusters from the functional enrichment analysis were used to generate a protein-protein interaction (PPI) network and visualised with Cytoscape^17^ (v4.01) using ranked gene set enrichment analysis (GSEA)^18^.

## Results

### COVID-19 severity signatures are enriched during subclinical and active tuberculosis

Nine COVID-19 studies with 35 signatures were eligible for evaluation (figure 1, table 1). Three TB cohort studies encompassing 853 individuals and 1181 samples, at various time points, were eligible for comparison. COVD-19 signatures were first evaluated in the TB combined observational and prospective cohort (TCC), from the UK (London and Leicester) and South Africa, including 293 individuals^19^. Out of the 35 COVID-19 signatures profiled, 25 were significantly associated (p<0·005) with higher COVID-19 risk scores in TB progressors and ATB patients, compared with latently infected individuals. Conversely, the influenza signature centred on a score of zero, across the spectrum of TB infection in both countries (figure 2). Of the ten COVID-19 signatures that were not associated with a significantly higher COVID-19 risk score, across the TB spectrum, seven were enriched in those with mild COVID-19 in the original studies compared to severe COVID-19 patients. Of the 20 scRNA-seq immune cell population signatures profiled; innate immune cell signatures generated the highest COVID-19 risk scores in the ATB groups. The classical monocyte signature^20^ was drastically increased in ICU cases of COVID-19, yielding the most significantly increased risk scores between LTBI and ATB samples (p<0·0001). The Silvin *et al*. COVID-19 WB neutrophil signature^21^ was most significantly increased in ATB vs LTBI (p<0·0001). The lung macrophage sub-populations associated with severe disease in BALF from COVID-19 patients^13^ showed high risk scores; in particular, the *FCN1*^hi^ (monocyte-derived macrophages (MDMs), G1), *FCN*^lo^*SPP1*^hi^ (pre-fibrotic macrophages, G2), and intermediary G1/2 macrophages. Conversely, the majority of adaptive immune cell signatures (CD4 and CD8 T cells) were higher in LTBI and TB contacts who didn’t progress to TB (figure 2). In general, T cell populations are depleted during severe COVID-19 infection^13,20,22^ and a lower COVID-19 risk scores for these signatures in the active and progressive TB patients could reflect a similar depletion of these populations. In the Wei *et al*. study of patients recovering from COVID-19^20^, the monocyte signature was associated with the early recovery stage indicating persistence of hyperinflammatory response, whereas the NK cell, T cell and B cell signatures were enriched in the late recovery stage, explaining the low COVID-19 risk scores associated with these signatures in ATB cases.

**Figure 2.**
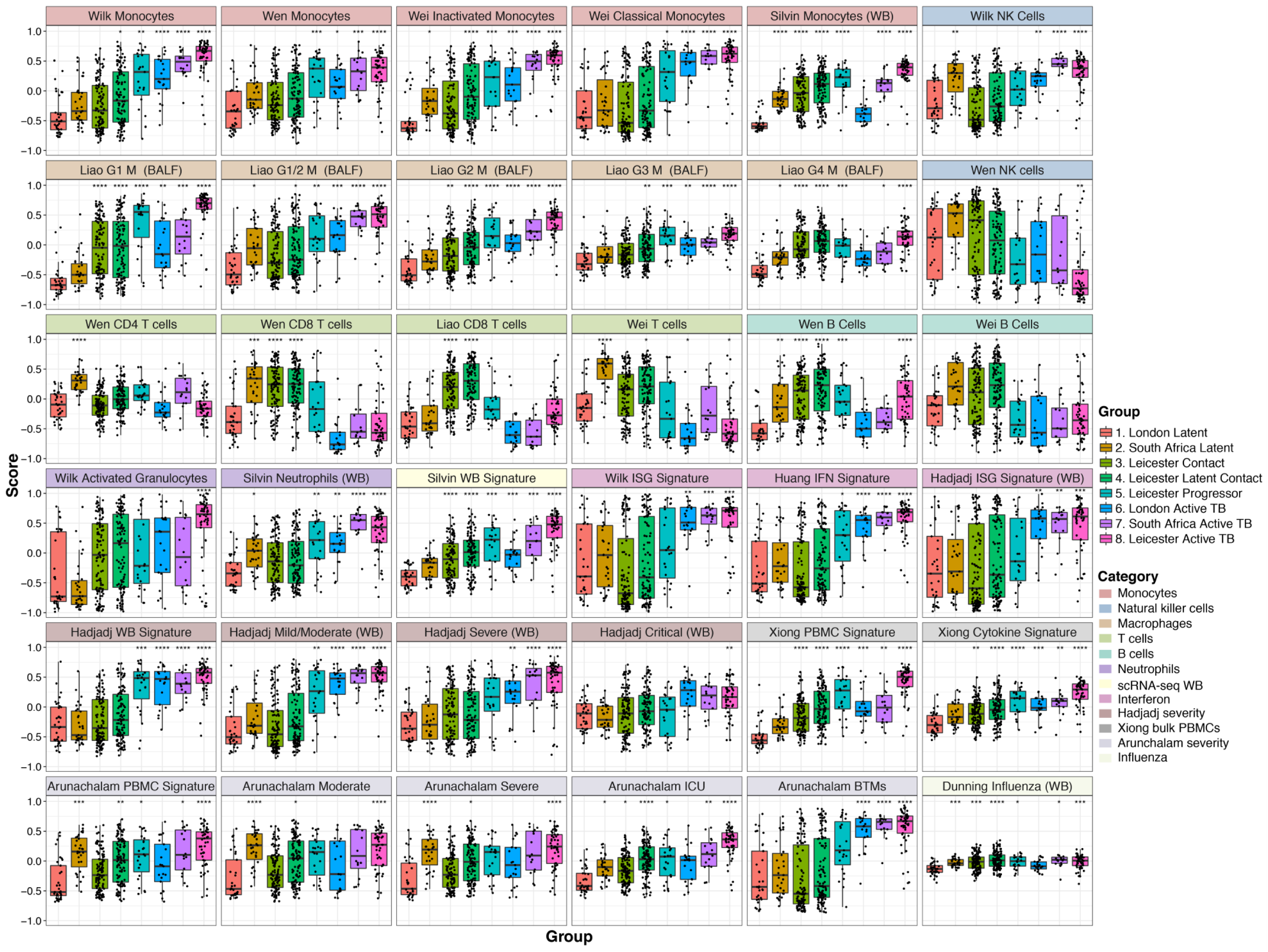
Profiling immune cell signatures from COVID-19 patients highlights increasing risk of severe disease associated with progression to active tuberculosis. COVID-19 immune cell signatures were derived from bulk and single-cell RNA-sequencing (RNA-seq) studies and used to generate putative “COVID-19 risk scores” from a tuberculosis (TB) whole blood bulk RNA-seq dataset using the TBSignatureProfiler package. TB samples are grouped according to disease state and COVID-19 signatures were categorised by immune cell or signature type. Scores for each signature were compared by contrasting each group with the London Latent group using a t-test adjusted for multiple testing using a Bonferroni correction. All signatures were derived from peripheral blood mononuclear cells (PBMCs) unless otherwise stated in the boxplot title. WB, whole blood; Mφ, macrophage; BALF, bronchoalveolar lavage fluid; ISG, interferon (IFN)-stimulated gene; NK, natural killer; ICU, intensive care unit; BTM, blood transcriptional module. *<0·05, **<0·005, ***<0·0005, ****<0·00005.

The mild/moderate and severe COVID-19 WB signatures^23^ (figure 2, Hadjadj *et al*.) showed significant differences in COVID-19 risk scores between ATB and LTBI (p<0·00001 and p<0·005, respectively) while the critical signature generated a low COVID-19 risk score which was significantly higher in the Leicester ATB cases than the LTBI cases (p<0·0001). COVID-19 disease severity blood transcriptional module (BTM) signatures^24^ included genes involved in IFN responses and antigen processing and presentation and clearly separated active disease samples from LTBI and QFT-negative TB contact samples (P<0·0001).

Assessing COVID-19 risk scores after further stratifying the Leicester TB contacts and ATB index cases by the location of index case TB (pulmonary [PTB] or extrapulmonary [EPTB]) revealed higher COVID-19 risk scores in contacts of PTB and PTB patients than those of EPTB, respectively (appendix 1 p6).

### COVID-19 signatures separate latent tuberculosis cases from those who progress to active disease by COVID-19 risk score

Thirteen signatures having significantly higher COVID-19 risk score (with an associated adjusted p<0·005) in at least two ATB groups, compared with LTBI, were selected for validation in the additional two TB progressor RNA-seq datasets: the ACS^25^ and GC6 cohorts including 153 and 407 individuals, respectively.^26^ Both datasets exhibited the same trend of increased COVID-19 risk score for all 13 signatures in progressors compared with LTBI (p≤0·01), whilst scoring zero for the influenza signature. The COVID-19 IFN signatures^23,27,28^ were associated with the greatest difference in COVID-19 risk score between LTBI and progressor groups, p<0·001 for both datasets). Plotting by days to TB diagnosis revealed an additional trend of higher risk of severe COVID-19 associated with proximity to ATB disease in the ACS cohort (figure 3A), but not the GC6 cohort (figure 3B).

**Figure 3.**
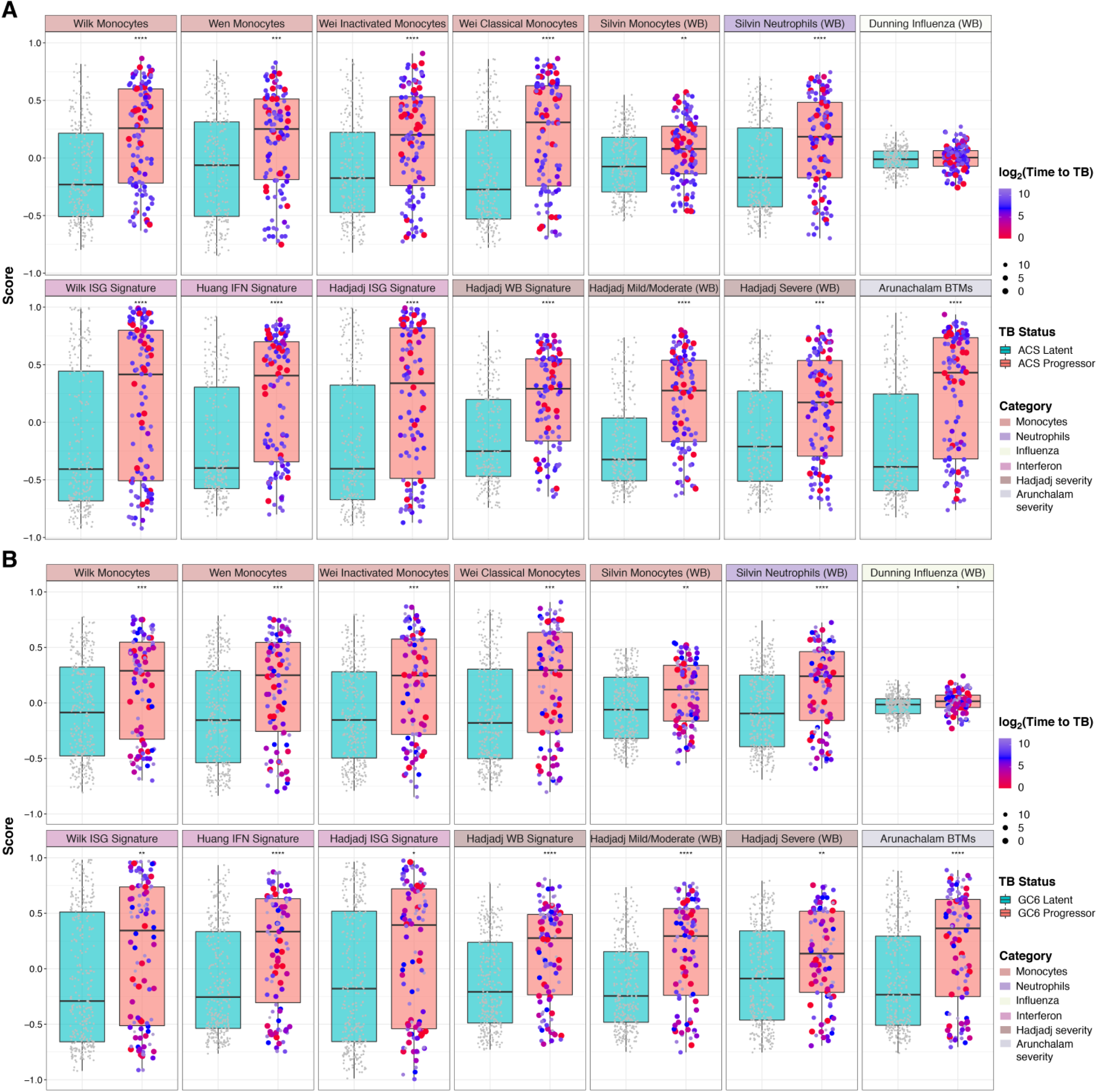
Risk of developing severe COVID-19 is significantly elevated in patients that progress from latent to active tuberculosis disease. COVID-19 immune cell signatures that were associated with significant differences in COVID-19 risk score between controls and progressor/active tuberculosis (TB) cases were validated on two additional whole blood RNA-sequencing TB datasets - (A) the Adolescent Cohort Study and (B) the Grand Challenges 6 study. TB samples were classified as latent or progressors and COVID-19 signatures were categorised by immune cell or signature type. Samples from patients that progressed to active TB disease during the study follow-up period are coloured and scaled according to time to TB diagnosis, measured in days and plotted on a log2 scale. Scores for each signature were compared by contrasting progressor with latent cases using a t-test adjusted for multiple testing using a Bonferroni correction. All signatures were derived from whole blood unless otherwise stated in the boxplot title. *<0·05, **<0·005, ***<0·0005, ****<0·00005.

### High concordance between monocyte subpopulations identified in BALF of severe COVID-19 patients and those in circulation during active TB disease

Reported similarities between BALF and WB scRNA-seq expression profiles observed in COVID-19 patients led us to investigate whether the BALF macrophage sub-lineages^13^ that associated with COVID-19 risk in ATB patients (figure 2) could be detected in circulation during TB infection. An integrated scRNA-seq analysis was performed using publicly available TB scRNA-seq PBMC data, consisting of both active and LTBI samples with BALF scRNA-seq data^13^, from COVID-19 patients of varying disease severity (appendix 1 p7). A high concordance was observed between the immune cell populations present within ATB PBMC and severe COVID-19 BALF after tSNE dimensionality reduction (figure 4A, left panel). Canonical cell type marker genes were identified for each cluster and assigned to the tSNE plot to identify shared and unique subpopulations (figure 4A, right panel). Three major macrophage sub-lineage markers identified in the original BALF scRNA-seq analysis^13^, *FCN1, SPP1* and *FABP4*, were separately profiled on the macrophage clusters of the COVID-19 and TB samples. Of the three markers, *FCN1* had highest expression in the TB PBMCs, while *FABP4* was completely absent. The *FCN1*-expressing pro-inflammatory monocyte-derived macrophage population was the most abundant of the sub-lineages in severe COVID-19 patients^13^. Zooming in on the *FCN1*- and *SPP1*-expressing clusters, the additional inflammatory markers that were identified for these populations in the original analysis were profiled. Both ATB and severe COVID-19 samples had high expression of these markers (figure 4C) indicating that these immune cell sub-lineages are active in the inflammatory response elicited by both diseases in the lungs and the blood.

**Figure 4.**
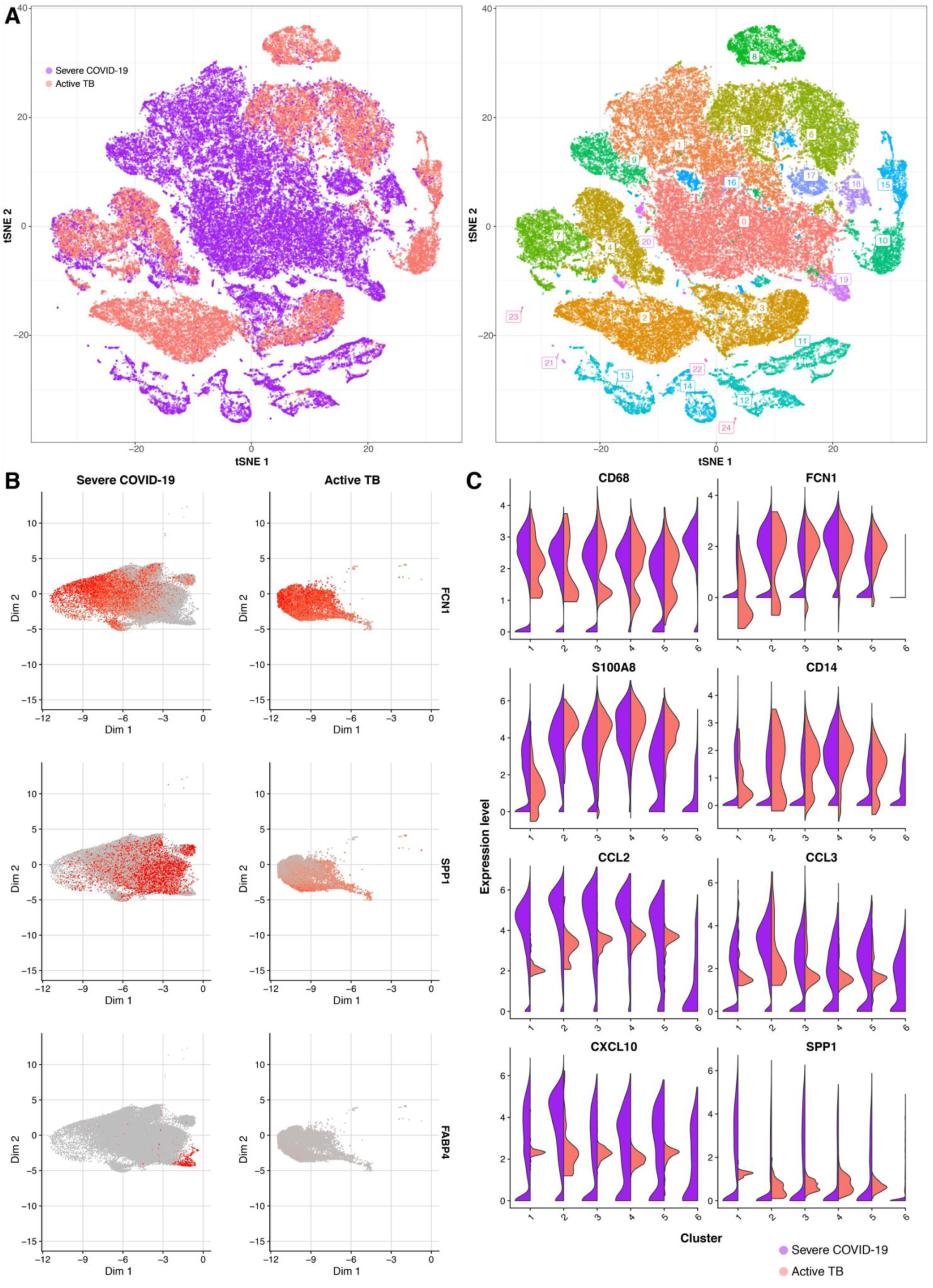
Macrophage subpopulations upregulated in the lungs of severe COVID-19 patients can also be found in the circulation during active tuberculosis disease. Single-cell RNA-sequencing (scRNA-seq) data from bronchoalveolar lavage fluid (BALF) of severe COVID-19 patients (n = 6) and peripheral blood mononuclear cells (PBMCs) from patients with active tuberculosis (TB) disease (n = 3) were integrated. **(A)** t-distributed stochastic neighbour embedding (tSNE) plot of integrated scRNA-seq data in the left panel, with cells from severe COVID-19 patients in guava and those from active TB patients in cyan, with corresponding cell clusters annotated based on identified markers in the right panel. **(B)** Macrophage clusters from the severe COVID-19 patients (left column) and active TB patients (right column), with the expression of major macrophage subpopulation markers identified in the original COVID-19 study - *FCN1*, high in G1, low in G2; *SPP1*, G2 and G3; *FABP4*, G4 - highlighted in red. **(C)** Violin plots depicting the expression levels of additional inflammatory marker genes associated with the macrophage subpopulations also present in the TB PBMC data, for each macrophage cluster.

### Highly similar enriched ontologies are shared between COVID-19 and tuberculosis

A meta-pathway enrichment analysis was performed using the transcriptomic data from the Leicester TB^19^, the COVID-19 WB scRNA-seq data^21^, and the influenza viral control cohort^9^. Among the top 1000 DEGs (selection explained in appendix 1 p3,8), DEGs from COVID-19, ATB and TB progressors were enriched for similar pathways compared to LTBI and influenza (figure 5A, appendix 1 p8, appendix 2 p5). Among the top 100 pathways across all groups, the highest percentage was associated with COVID-19 (96% of those considered), followed by ATB (93%), progressors (88%), influenza (64%), and LTBI (28%) (figure 5B). Comparing the twenty most significant ontologies, influenza and COVID-19 patients could be distinguished on the basis of an absence of IFN-γ response, lack of TNF signalling, both of which were highly enriched in TB progressors and COVID-19 (figure 5A). Interestingly, LTBI had no enrichment of cytokine production and regulation of innate immune response compared with the other disease states, indicating a less active immunological response (figure 5A).

**Figure 5.**
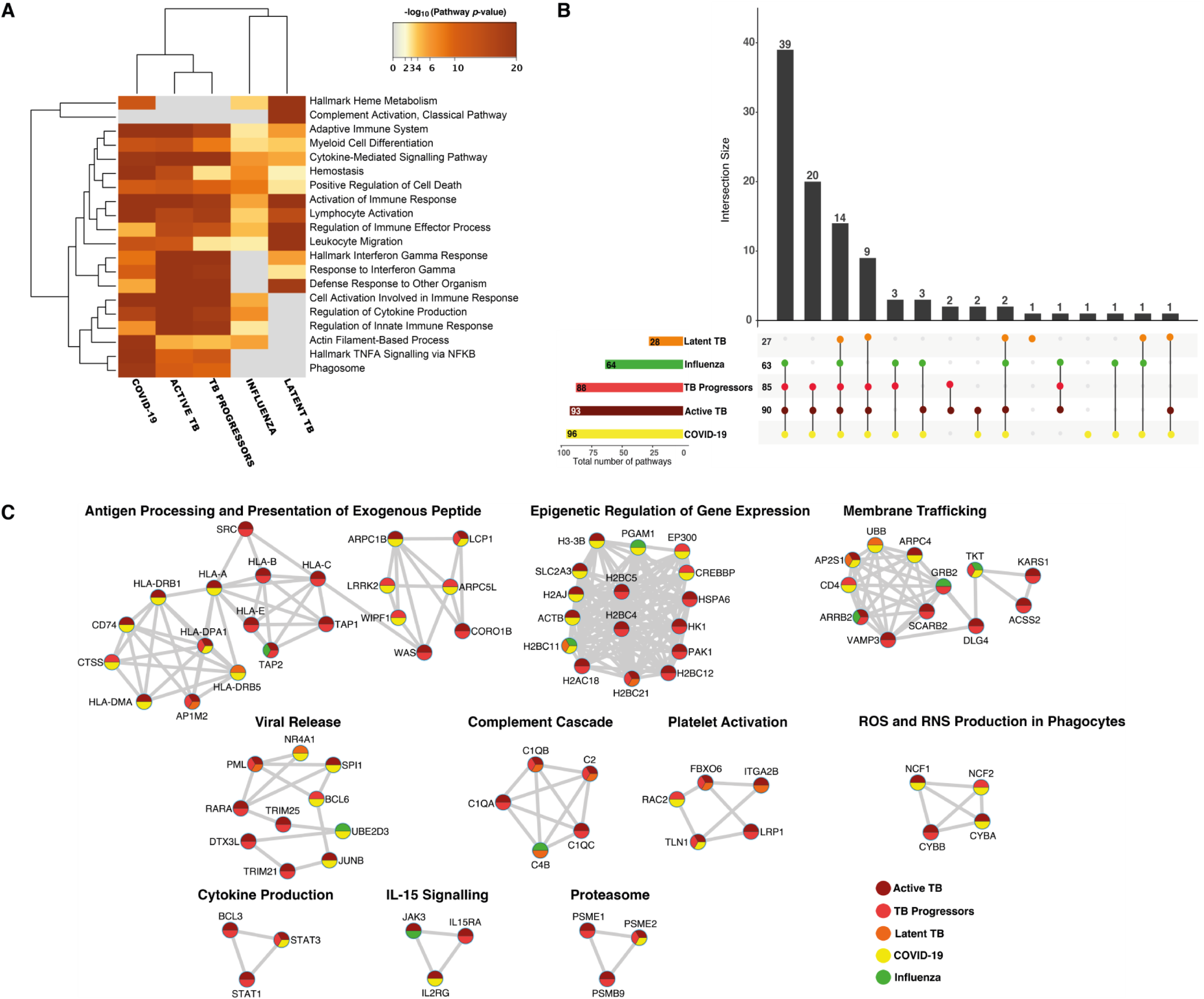
Active TB disease and COVID-19 have similar perturbed pathways and gene networks. **(A)** Heatmap of the top 20 enriched pathways from the meta-analysis across the groups with the best p-values. Cells are coloured by their -log_10_(p-values), grey cells indicate a lack of significant enrichment for that term/ontology in the corresponding DEGs list. **(B)** Upset intersection plot showing the number of pathways shared between COVID-19 and the other groups compared. Wherever the genes are shared the specific coloured dots appear below the column bar graph which show the number of shared pathways, which are connected by vertical lines, denoting shared categories. The horizontal bars represent the category gene count and the numbers on the horizontal axes represent the common pathways between the represented category and COVID-19. **(C)** All MCODE components from a protein-protein interaction (PPI) network analysis of all merged gene lists are displayed as networks, nodes are displayed as pie charts, colour coded by disease group. The labels are added manually and are derived from functional labels based on the top three enriched terms for that cluster.

To identify some key effectors responsible for the activation of these pathways, ten significant protein-protein interaction (PPI) network clusters were identified and are represented as annotated networks in figure 5C. Among the 85 DEGs which were represented in the network analysis, only nine belonged to influenza, while there was greater alignment of PPI networks between COVID-19 and the TB groups, further confirming the similarity between COVID-19 and TB. Antigen processing and presentation was the largest cluster and was enriched mostly in COVID-19 and TB groups, with only one member gene representing influenza. The second most enriched cluster shared between COVID-19 and TB, annotated as epigenetic regulation of gene expression, consisted of many histone related proteins, suggesting both result in widespread changes in the epigenome. Other enriched clusters included platelet activation, cytokine production, and proteasome were exclusively enriched in COVID-19 and TB, indicating extensive commonality between perturbed pathways (figure 5C, p-values in appendix 2 p6).

### Genes clustered based on similarity between COVID-19 and tuberculosis associate with COVID-19 disease severity

Three clusters, based on similarity between the other diseases and COVID-19, were identified (figure 6A). Cluster 1: common to all TB disease states, but absent from influenza, consists of IFN-γ response, apoptosis signalling and B cell activation pathway enrichment. Cluster 2: common across all disease groups, except for LTBI, consists of complement activation, inflammatory response, apoptotic signalling, T cell receptor signalling, and IFN-γ production. Cluster 3: shared between ATB and TB progressors, but absent from LTBI and influenza, was enriched for platelet degranulation, autophagy, antigen processing and presentation, TNFA signalling, IL6/JAK/STAT3 signalling, and cellular processes indicating higher metabolic requirements of cells in response to infection. Regulation of mTORC1 signalling was the only pathway exclusively enriched in COVID-19 from the top 100 enriched pathways (figure 6A). To further explore if the enriched pathway DEGs were associated with severity of COVID-19, GSEA was used to determine enrichment of the three identified clusters in a WB bulk RNA-seq dataset containing samples from different clinical stages of COVID-19^21^. Cluster 1, was found to be enriched in all three COVID-19 disease severity categories with moderate effect size, while cluster 2 and cluster 3 were enriched in more severe disease states with larger effect sizes as indicated by the *q*-values in figure 6B. DEGs common to COVID-19 and TB states were significantly enriched with increasing COVID-19 severity; 17% were enriched in moderate COVID-19, 18% were enriched in severe COVID-19 and the greatest proportion, 32%, were enriched in the ICU group (appendix 1 p9).

**Figure 6.**
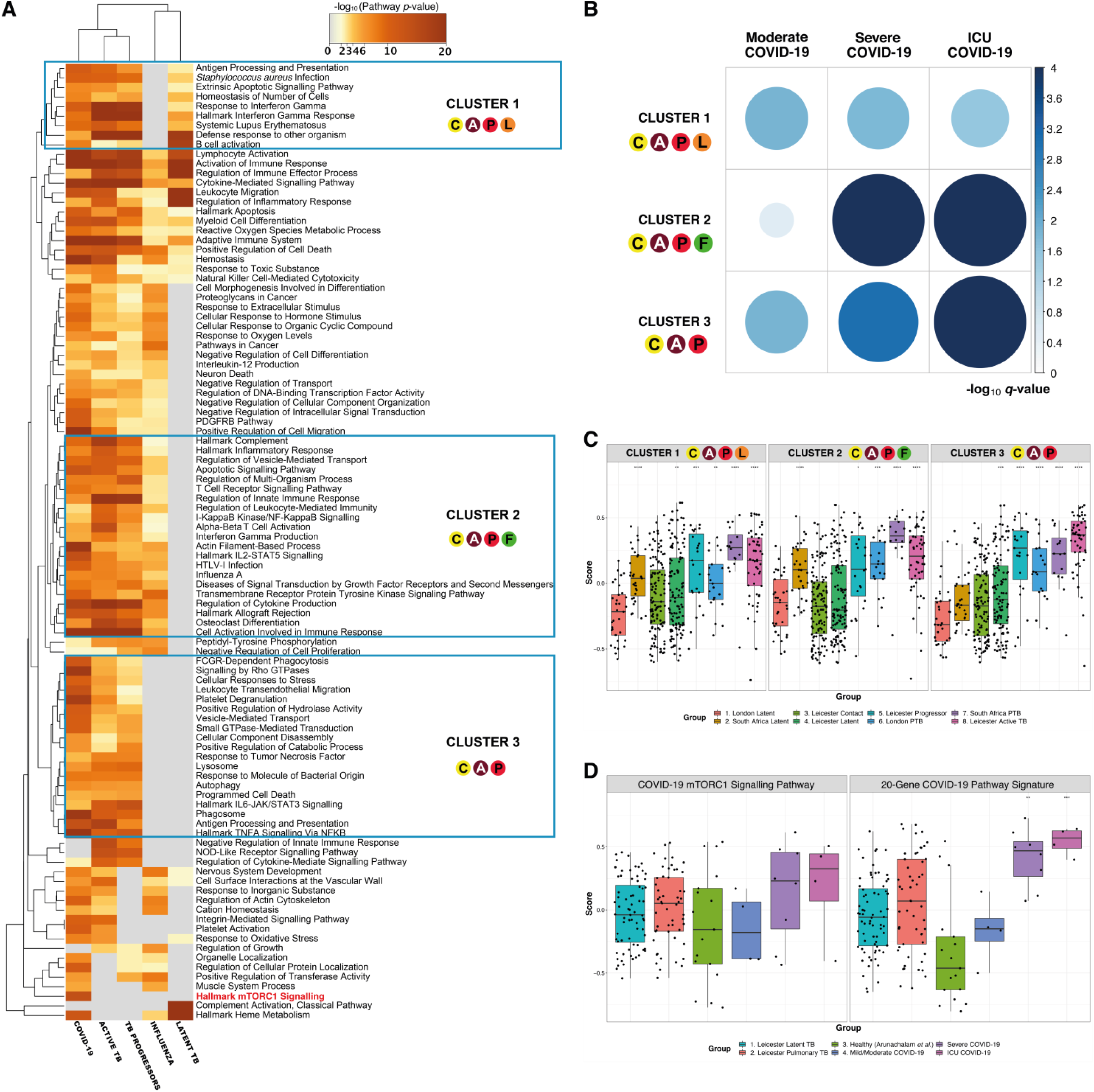
Shared COVID-19 and TB cluster differentially expressed genes correlate with disease severity and interfere with COVID-19-specific gene signature detection. **(A)** Heatmap of the top 100 enriched pathways and three identified gene clusters, the circle colours correspond to the groups that formed part of the cluster, the letters in the circle indicate the groups: C, COVID-19; A, active TB; P, TB progressors; L, latent TB; F, influenza. **(B)** Bubble plot depicting the enrichment of identified cluster differentially expressed genes (DEGs) by COVID-19 disease severity (appendix 2 p8). The size and shade of the circle correspond to the -log10 *q*-value, represented as a scale on the right. **(C)** Box plots of the gene set variation analysis (GSVA) of COVID-19 scores based on the DEGs of each of the three identified clusters. Membership of the groups in the cluster is indicated in the circle appearing in the title of each boxplot. Scores for each signature were compared between each group at the “London Latent” control group using a t-test adjusted for multiple testing using a Bonferroni correction. **(D)** All pathways identified in the pathway meta-analysis were considered and selected based on COVID-19 exclusivity; bar plot of GSVA for mTORC1 signalling pathway genes and the reduced COVID-19 20-gene signature, when profiled on the Leicester cohort and COVID-19 disease severity data. Scores for each signature were compared between each group at the “Healthy (Arunachalam *et al*.)” control group using a t-test adjusted for multiple testing using a Bonferroni correction. *<0·05, **<0·005, ***<0·0005,****<0·00005.

The overlap between DEGs associated with COVID-19 disease severity and TB raised the possibility of an impediment to the discovery of a specific COVID-19 diagnostic WB biomarker. TBSignatureProfiler was run on cluster-specific DEGs using TCC data. In all three clusters, groups ranging from TB progressors to ATB showed significant COVID-19 risk scores compared with the LTBI control group (figure 6C). Cluster 3, which contains the most shared DEGs between COVID-19, ATB and TB progressors, exhibited the most graded significant increase in COVID-19 risk scores among the three clusters, suggesting that shared DEGs between COVID-19 and TB would interfere with the derivation of a COVID-19-specific diagnostic signature (figure 6C, appendix 2 p9).

To determine whether a more specific COVID-19 diagnostic signature could be identified what would not be affected by TB co-infection, DEGs from the only COVID-19-specific pathway, mTORC1 signalling (figure 6C), were used to compare the COVID-19 risk scores between the Leicester TB cohort and the COVID-19 disease severity data. The mTORC1 signature demonstrated a clear increase in COVID-19 risk score with increasing COVID-19 disease severity (figure 6D); lower COVID-19 risk scores were generated from the ATB group, with the LTBI contacts and the healthy controls having similar scores. Using the full list of 820 exclusive pathways enriched in COVID-19 (appendix 2 p10) a 20-gene signature was derived by selecting genes from the top 10 significant pathways and excluding DEGs shared between any other disease group, enriched for significance using a ranked GSEA on the COVID-19 severity data (appendix 1 p10–11). This reduced signature yielded significant differences in risk scores between healthy controls and severe COVID-19 (p<0·005) and ICU patients (p<0·0005), and not TB groups (figure 6D, appendix 2 p13).

## Discussion

Emerging findings to aid rapid responses to the COVID-19 pandemic has led to an increased deposition of manuscripts on pre-print servers. Whilst they require rigorous peer-review, they have served as a valuable resource for further hypothesis generation and testing. We used several such studies to augment investigation of our hypothesis using these sequencing data in combination with new bioinformatics packages developed for assessing TB diagnostic signature performance.^10,12^ Substituting TB signatures for COVID-19 signatures, within the TBSignatureProfiler package, we demonstrate the overlap between severe COVID-19 transcriptional signatures and WB TB disease phenotypes found in ATB patients and importantly those with asymptomatic TB who progress to symptomatic TB over 1–2 years.

The most compelling overlaps between COVID-19 signatures and the TB datasets reside in the circulating innate immune cells. Classical monocyte and neutrophil signatures derived from severe COVID-19 patients were associated with the highest COVID-19 risk scores when profiled across the TB spectrum. Circulating monocyte activation status is a determining factor for COVID-19 prognosis, with specific phenotypes leading to poorer outcomes^29^. Similar monocyte phenotypes have been detected during TB infection^30^ and the presence of these in circulation prior to SARS-CoV-2 co-infection may be detrimental to the activation of key adaptive antiviral immune responses.

Conversely, adaptive immune cell populations enriched in milder COVID-19 cases were associated with lower risk scores in ATB and progressors. Both T and B cells were significantly reduced in COVID-19 ICU patient samples compared to pre and post ICU^20^. Impairment of functional CD4^+^ and CD8^+^ T cells has been associated with severe COVID-19^31^ and may be exacerbated by the presence of exhausted T cell phenotypes indicative of chronic *Mtb* infection which display reduced capacity to produce effector cytokines^32^.

SARS-CoV-2-infected macrophages are known to accumulate in lungs of patients who died from COVID-19^33^. Activated macrophages play a major role in chronic inflammation in TB and HIV and increase the likelihood of severe COVID-19 infection (odds ratios: 1·7 and 2·3, respectively)^34^. *FCN1*^hi^ macrophages are abundant in the BALF of severe COVID-19 patients^13^ and in BALF indicating their migration to the lungs from the blood, substantiated by an integrative analysis between COVID-19 blood and BALF scRNA-seq samples performed by Silvin *et al*.^21^. We show that both the *FNC1*^hi^ and *FCN1*^lo^*SPP1*^*+*^ sub-lineages associated with significantly higher COVID-19 risk scores in recent contacts of TB patients, progressors, and ATB patients. Our integrative analysis of TB PBMC and COVID-19 BALF scRNA-seq samples showed the presence of these phenotypes in circulation during ATB, indicating that the presence of these sub-lineages in the blood may predispose TB patients to more severe lung inflammation.

The importance of neutrophils has largely been underestimated in existing scRNA-seq workflows, due to technical issues associated with the preservation and sequencing of neutrophils, despite evidence that neutrophils are correlated with poor prognosis and disease severity in COVID-19^21,35,36^, and out-of-control inflammation or a failure to establish adequate adaptive immune responses in severe TB^37^. The upregulation of calprotectin subunit genes *S100A8* and *S100A9* in these neutrophils may account for or trigger the cytokine storm that is characteristic of severe COVID-19^21^ and S100A8/A9-dependent neutrophilic accumulation in the lungs of ATB patients leads to induction of inflammatory mediators and promotes lymphocyte trafficking^38^. These findings underscore the need for more WB single-cell studies rather than PBMCs in order to characterise the important contribution of neutrophils to severe COVID-19.

IFN-induced transcriptional signatures were among the most significantly upregulated in severe disease in the studies selected risk profiling^20,24,27,28^ and generated high COVID-19 risk scores among ATB cases. Dysregulation of IFN production^23,39^ and the nature of type I and III IFN responses (location, timing, and duration) in COVID-19^40,41^ and TB^42^ guide disease progression and outcomes. It is therefore plausible that dysregulation of type I IFN responses during SARS-CoV-2 co-infection may also have an impact on TB disease progression.

At a systems levels, shared biological pathways showed a graded enrichment of similar pathways between COVID-19 and TB, particularly ATB. A PPI-based network analysis identified common genes, highlighting that shared molecular determinants between COVID-19 and TB disease states can influence the clinical outcome if both diseases affect the same individual i.e., during co-infection. Common enriched pathways between COVID-19 and TB signify activation of innate immune responses directed against both *Mtb* and SARS-CoV-2 and include antigen presentation^27,43^, membrane trafficking^44,45^, ROS/RNS production^46,47^, activation of complement^48,49^, cytokine production^50,51^, and platelet activation^52,53^. This suggests that both hyperactivation of these responses or evasion by the pathogen may lead to severe clinical presentations for both of these infections and be synergistically exacerbated in co-infection. These mechanistic similarities would complicate efforts to derive a COVID-19-specific transcriptional signature. We demonstrated this with a 20-gene COVID-19-specific signature that gave rise to significantly higher scores among severe COVID-19 cases compared with healthy controls, but also produced slightly elevated scores in TB-infected individuals.

Although several of the pre-print manuscripts identified at the beginning of our study have since been published in high-impact journals, our COVID-19 signature search was complicated by the limited availability of comprehensive supplementary data detailing cluster markers or DEG lists. Furthermore, the COVID-19 studies performed to date are confined to small numbers of patients in each group that exhibit inter-individual variability in immune cell phenotypes. We counteracted this limitation by including signatures from multiple studies for each immune cell population. In summary, we show for the first time through large scale meta-analysis of the available transcriptomic data, that advanced COVID-19 and TB disease states overlap at the gene, cell, and systems levels. These shared disease mechanisms could prove to be hotspots for immune exacerbation, inducing greater immunopathology, in the case of co-infection. We report a new 20-gene gene signature which distinguishes severe COVID-19 from active and LTBI that should be investigated further for its disease classification value in larger datasets as they become available. Taken together, the data presented here along with the emerging case reports identifying TB as a risk factor for severe COVID-19 suggesting that individuals with known previous TB history, recent TB exposures or LTBI with pre-existing lung pathology, are at increased risk of severe COVID-19 disease and, potentially, early progression to TB disease, following SARS-CoV-2 infection. Given the medical capacity to do so, we therefore propose that such individuals should 1) be closely followed to allow early detection of respiratory symptom onset, 2) be screened for SARS-CoV-2 and TB at symptom onset, and 3) be followed up for TB in the months subsequent to SARS-CoV-2 diagnosis. Diagnostic and clinical outcome data arising from early stages of the COVID-19 outbreak should be stratified by TB history in order to determine the effects of these co-infections as soon as possible.

## Data Availability

All data used in the present study are currently publicly available and their sources are referenced in the main text of the manuscript. R scripts used for the analysis will be made available on GitHub when the manuscript has been submitted to a journal for peer review. All R packages used in the analysis are publicly available and referenced.

## Declarations

### Acknowledgments

The authors would like to thank colleagues at the Wellcome Centre for Infectious Diseases in Africa and the Walter and Eliza Hall Institute of Medical Research for their feedback throughout the study.

### Authors’ contributions

The study was conceived and designed by AC, DS, and A. DS and A contributed equally to the data analysis. AC supervised the analysis and assisted with the study design. Systematic review of COVID-19 manuscripts and identification of TB datasets was performed by DS and A. The curatedTBData package used to obtain the TB transcriptomic data and associated patient metadata was created by XW and WEJ. COVID-19 signature risk profiling and scRNA-seq analysis was performed by DS with input from WEJ, who developed the TBSignatureProfiler package with colleagues. Functional enrichment analysis and GSEA were performed by A. DS, A, and AC prepared the manuscript, with editorial input from all authors.

